# HDL-bound microRNAs modulating cholesterol efflux and homeostasis and incidence of acute myocardial infarction: A population-based case-cohort study

**DOI:** 10.64898/2026.02.11.26346068

**Authors:** Javier Hernando-Redondo, Mariona Llaves, Álvaro Hernáez, Pau Berenguer, Daniel Muñoz-Aguayo, Marta Bódalo, Julia Perera, Noemí Rotllan, Joan Carles Escolà-Gil, Dolores Corella, Olha Khymenets, Anna Camps-Vilaró, Isaac Subirana, Jaume Marrugat, Roberto Elosua, Albert Goday, Lluis Recasens, Mary Cano-Sarabia, Daniel Maspoch, Marta H. Hernández, Irene Roman-Degano, Aleix Sala-Vila, Iolanda Lázaro, Olga Castañer, Montserrat Fitó

**Affiliations:** Registre Gironí del Cor (REGICOR) Study Group, Hospital del Mar Research Institute, 08003 Barcelona, Spain; Cardiovascular Risk and Nutrition Research Group, Hospital del Mar Research Institute, 08003 Barcelona, Spain; Consorcio CIBER, M.P. de Fisiopatología de la Obesidad y Nutrición (CIBEROBN), Instituto de Salud Carlos III, 28029 Madrid, Spain; Ph.D. Program in Food Science and Nutrition, University of Barcelona, Barcelona, Spain; Universitat Ramon Llull, Blanquerna School of Health Sciences, 08025 Barcelona, Spain; Consorcio CIBER, M.P. de Enfermedades Cardiovasculares (CIBERCV), Instituto de Salud Carlos III, 28029 Madrid, Spain; Bioinformatics Unit, MARData, Hospital del Mar Research Institute, 08003 Barcelona, Spain; MARGenomics, Hospital del Mar Research Institute, 08003 Barcelona, Spain; Institut de Recerca Sant Pau, 08041 Barcelona, Spain; Consorcio CIBER, M.P. de Diabetes y Enfermedades Metabólicas Asociadas (CIBERDEM), Instituto de Salud Carlos III, 28029 Madrid, Spain; Department of Preventive Medicine, University of Valencia, 46100 Valencia, Spain; Applied Metabolomics Research Group, Neurosciences Research Programme, Hospital del Mar Research Institute, 08003 Barcelona, Spain; Faculty of Medicine, University of Vic - Central University of Catalunya, 08500 Vic, Spain; Cardiovascular Epidemiology and Genetics Research Group, Hospital del Mar Research Institute, 08003 Barcelona, Spain; Department of Endocrinology and Nutrition, Hospital del Mar, 08003 Barcelona, Spain; Department of Medicine, Universitat Autònoma de Barcelona, 08193 Cerdanyola del Vallès, Spain; Department of Cardiology, Hospital del Mar, 08003 Barcelona, Spain; Heart Diseases Biomedical Research Group, Hospital del Mar Research Institute, 08003 Barcelona, Spain; Catalan Institute of Nanoscience and Nanotechnology, Spanish National Research Council–Barcelona Institute of Science and Technology, 08193 Cerdanyola del Vallès, Spain; ICREA–Catalan Institution for Research and Advanced Studies, 08010 Barcelona, Spain; Consorcio CIBER, M.P. de Epidemiología y Salud Pública (CIBERESP), Instituto de Salud Carlos III, 28029 Madrid, Spain

**Keywords:** Atherosclerosis, HDL-miRNAs, Myocardial Infarction, Incidence, hsa-miR-628-3p, hsa-miR-28-3p

## Abstract

**Background:** HDL particles can carry microRNAs (miRNAs), capable of regulating gene expression connected to HDL functions. Despite links to some cardiovascular risk factors, miRNA association with incident acute myocardial infarction (AMI) remains unclear.

**Objectives:** Our aim was to elucidate the association between HDL-bound miRNAs (HDL-miRNAs) and incident AMI using a non-targeted approach in a population-based study.

**Methods:** We conducted a case-cohort study including 247 participants from the REGICOR cohort in northeastern Spain (51 AMI cases and a random sample of 196 participants, including seven overlapping AMI cases). HDL-miRNAs were isolated from apolipoprotein B-depleted serum and quantified by whole-genome miRNA sequencing. Associations between HDL-miRNAs and incident AMI were assessed using multivariable Cox proportional hazards model. For AMI-associated HDL-miRNAs (*p*-value <0.10), we retrieved their experimentally validated targets and assessed pathway enrichment of these targets via over-representation analysis.

**Results:** Two HDL-miRNAs were associated with incident AMI after FDR correction: miR-628-3p (HR 1.69, 95% CI 1.30 to 2.19) and miR-28-3p (HR 1.58, 95% CI 1.21 to 2.06). Nine additional HDL-miRNAs were nominally associated with AMI incidence (*p*-value <0.05), eight with a direct association (miR-93-5p, miR-26b-5p, miR-106a-5p, miR-126-3p, miR-15b-5p, let-7a-5p, let-7e-5p, and let-7f-5p) and one with an inverse association (miR-361-5p). These miRNAs regulate the expression of genes in pathways involved in cholesterol regulation, particularly cholesterol efflux and homeostasis. The AMI group exhibited higher variance and a greater number of significant and strong correlations.

**Conclusions:** Two HDL-miRNAs (miR-628-3p and miR-28-3p) were significantly associated with AMI incidence. A tighter coregulatory network in cases was observed, underscoring their potential clinical utility in risk prediction and cardiovascular prevention.

**Clinical Perspective:** *What Is New?:*

- In a population-based case-cohort study we profiled the HDL-bound miRNome and found two miRNAs (miR-628-3p and miR-28-3p) associated with incident AMI.
- The use of HDL-enriched serum fractions provided a focused analysis on HDL functionality. These miRNAs regulate the expression of genes in pathways involved in cholesterol efflux and homeostasis (*ABCA1, ARL4C, SIRT1, NFKBIA, ANXA2, LRP6*) and show a tighter coregulatory network among significant miRNAs among cases, supporting biological coherence.

*What Are the Clinical Implications?:*

- HDL-miRNA signatures may complement traditional risk factors to refine AMI risk stratification and provide a rationale for HDL-guided, miRNA-targeted preventive interventions using HDL-like delivery platforms.

## 1. INTRODUCTION

Although low high-density lipoprotein (HDL) cholesterol is epidemiologically linked to cardiovascular disease (CVD) and other chronic conditions, its concentration alone poorly reflects the diverse functions of HDL, highlighting the need to shift diagnostic and therapeutic strategies toward optimizing HDL functionality[1]. HDL functions include cholesterol efflux capacity (the HDL ability to remove cholesterol excess from peripheral cells such as macrophages), as well as anti-inflammatory and antioxidant properties, which have been inversely associated with the incidence of coronary events [2–4] and all-cause mortality [4]. HDL functions depend on a number of proteins, bioactive lipids, and certain nucleic acids [1]. While the majority of HDL-associated micro RNAs (miRNAs) originate from longer RNAs with no clinical relevance, others regulate cell homeostasis or differentiation, apoptosis, and immune responses due to their capacity to modulate gene expression [5]. Circulating miRNAs are relevant to CVD, regulating endothelial cell differentiation, atherogenesis, and cholesterol efflux ^7^, and they have been studied as biomarkers of the incidence of coronary disease in prospective studies [8,9]. Nevertheless, few studies have investigated the role of nucleic acids related to HDL in CVD[10,11]. To our knowledge, no previous studies have approached HDL-miRNAs and incidence of cardiovascular or coronary events such as acute myocardial infarction (AMI). Therefore, our study aims to explore the relationship between the HDL-miRNome and AMI incidence in a well-characterized population-based cohort of Spanish adults using a non-targeted approach.

## 2. METHODS

### 2.1. Study design

We conducted a case-cohort study within the Registre Gironí del Cor (REGICOR) cohort, a representative sample of community-dwelling adults aged 35-75 years free of history of CVD at baseline. The full cohort comprised 5,404 individuals, 117 of whom developed coronary artery disease during follow-up [8]. From the case-cohort study extracted from the REGICOR cohort by Sanllorente et al [2], we randomly selected a subcohort of 196 individuals (including 7 overlapping AMI cases) and added 51 AMI cases, all with available biological samples (**Figure 1**). The median follow-up time for AMI cases was 4 years. Detailed information regarding the study protocol, recruitment strategies, inclusion/exclusion criteria, and data collection methods has been previously documented [12]. This study is presented following the Strengthening the Reporting of Observational Studies in Epidemiology guidelines.

**Figure 1.**
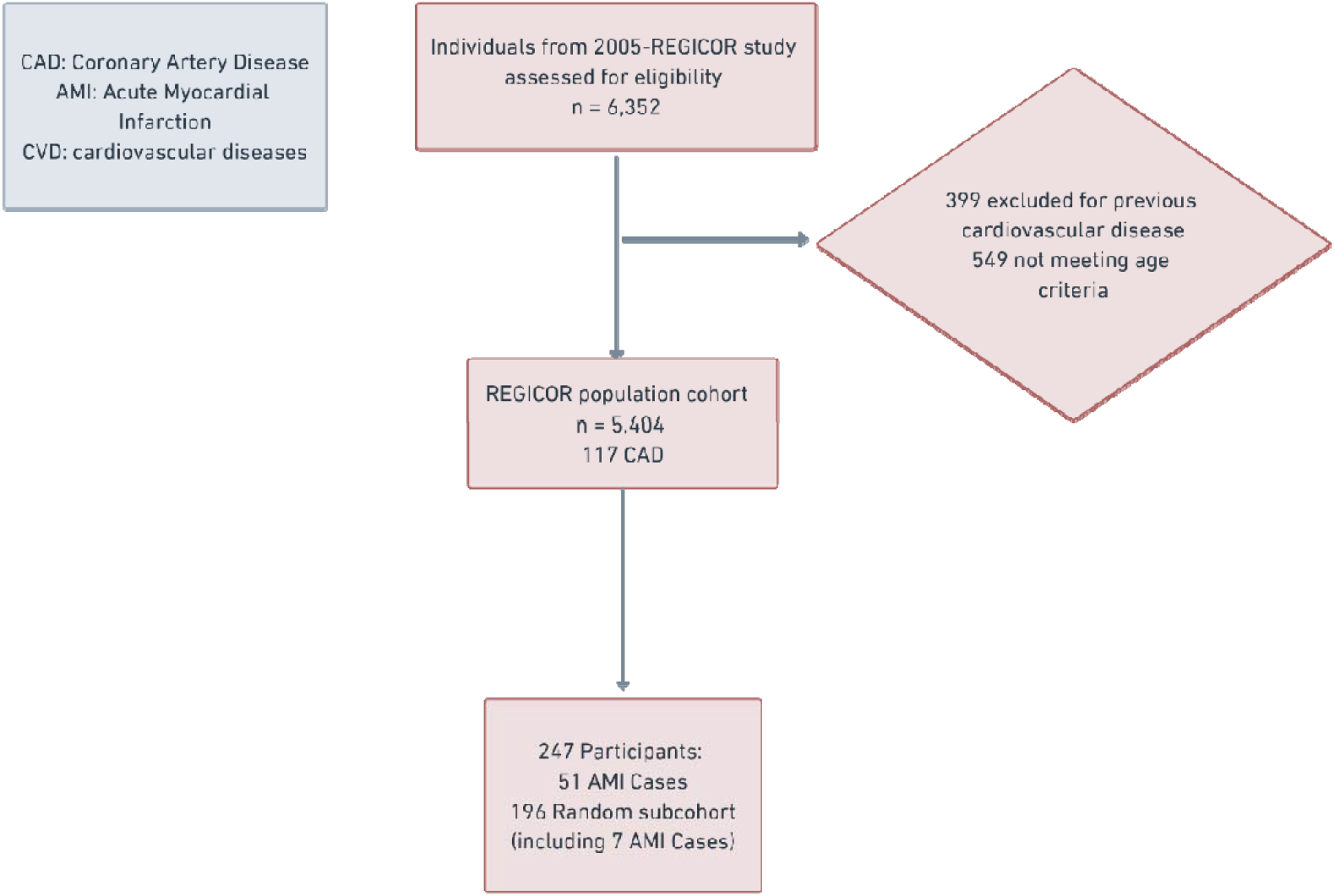
Flow chart of the study

### 2.2. Quantification of HDL-miRNAs

We obtained one fasting serum sample per participant, therefore each participant represents one biological replicate, and no technical replicates were performed. Samples were aliquoted and stored at −80°C until analysis. Apolipoprotein B-depleted serum samples, in which HDL are the only lipoproteins present, were prepared by precipitating apolipoprotein B-containing lipoproteins using 20% (w/v) polyethylene glycol 8000 (Sigma-Aldrich, Madrid, Spain) [13]. Total RNA, including miRNAs, was extracted from these samples using the Qiagen miRNeasy Serum/Plasma Advanced Kit (Qiagen, Hilden, Germany). We quantified HDL-miRNA expression using whole genome miRNA sequencing in a non-targeted approach, following the manufacturer’s protocol for serum/plasma samples (QIAseq miRNA Unique Dual Index Library Kit, HB-2983-001, November 2021). The kit incorporates Unique Molecular Identifiers to ensure accurate quantification of individual miRNA molecules. We sequenced the libraries on Illumina platforms, generating approximately 5 million reads per sample (75 bp, single-end). Lowly expressed miRNAs were removed to focus on robust signals, resulting in 111 HDL-miRNAs expression values in each sample, and data were normalized to account for differences in sequencing depth between samples. Quality control procedures identified the batch effect specific to one box which was subsequently accounted, and samples with insufficient read counts were excluded. Further technical details are provided in Supplementary Methods (Supplementary material).

### 2.3. Outcome: acute myocardial infarction

The endpoint was the fatal or non-fatal first occurrence of AMI confirmed by an expert clinical committee[8] following standardized guideline criteria [14]. Details of the AMI case investigation has been described elsewhere[15]. Non-fatal outcomes were determined through regular, standardized telephone interviews for up to 10 years combined with a review of medical records. Fatal outcomes were ascertained through linkage with the Public Data Analysis for Health Research and Innovation Program of Catalonia (PADRIS) of the Agency for Health Quality and Assessment of Catalonia (AQuAS) and National Statistics Institute. To avoid case duplication the information from all sources was linked and verified by the researchers.

### 2.4 Other variables

The following analytes were quantified in fasting serum samples using an ABX Pentra-400 auto-analyzer (Horiba-ABX, Montpellier, France): glucose (mg/dL), triglycerides (mg/dL), HDL cholesterol (mg/dL), and total cholesterol (mg/dL). LDL cholesterol was calculated using the Friedewald formula whenever triglycerides were below 300 mg/dL. Trained personnel collected information on clinical and lifestyle variables [12]. Body mass index was calculated as weight (measured using a calibrated scale) in kg divided by height (measured using a calibrated stadiometer) in meters squared. Hypertension was defined as systolic blood pressure ≥140 mmHg, diastolic blood pressure ≥90 mmHg, or the use of antihypertensive medication. Type 2 diabetes was defined as fasting glucose levels ≥126 mg/dL or the use of insulin or glucose-lowering medications. Hypercholesterolemia was defined as total cholesterol levels ≥200 mg/dL or the use of cholesterol-lowering medications. Regarding tobacco use, participants were categorized as never smokers, former smokers, or current smokers. Current smokers were defined as individuals who reported smoking regularly or having quit within the past year. Former smokers were those who had quit smoking more than one year ago. Never smokers were participants who reported never having smoked.

### 2.5. Ethical aspects

All procedures adhered to the principles of the Declaration of Helsinki. The study protocol was approved by the Hospital del Mar Clinical Research Ethics Committee (reference: 2015/6202/I). Prior to participation, individuals provided written informed consent.

### 2.6. Sample size

A sample size of 182 individuals from a sub-cohort drawn from a cohort composed of 5,404 participants in a case-cohort design allows ≥80% power to detect a hazard ratio (HR) of ≥2.25 or ≤0.44 when comparing HDL-miRNA expression in participants in the third tertile to those in the first tertile, assuming a 2-sided type I error of 5% and a 10-year incidence of the outcome of 1% [16].

### 2.7. Statistical analysis

Normally distributed continuous variables were described with means and standard deviations (SD), non-normally distributed continuous variables with medians and first-third quartiles, and categorical variables with proportions. HDL-miRNA levels were transformed into standardized units.

We calculated HRs for each one-SD increase in HDL-miRNA levels using Cox proportional hazards regression models considering the case-cohort design and the “LinYing” adjustment method. Age was used as the underlying time scale, with entry time defined as the participant’s age at recruitment and exit time as the age at the event, or the censoring date (whichever occurred first). Models were adjusted for: age, sex, technical batch, HDL cholesterol, total cholesterol, body mass index, diabetes, hypertension, and smoking. Adjustments for multiple comparisons (*n* = 111) were performed using the False Discovery Rate.

We explored expression structure among the HDL-miRNAs associated with incident AMI (*p*-value <0.10). We ran a principal component analysis in AMI cases and controls separately to evaluate which individual miRNAs contributed most to variance within each group. To examine inter-miRNA relationships, we computed pairwise correlation matrices among the selected miRNAs separately in AMI cases and controls (significant correlations were defined after multiple testing using the false discovery rate). Analyses were performed in R v3.5.2.

### 2.8. Functional analysis

We retrieved experimentally validated miRNA-target interactions among HDL-miRNAs linked to AMI incidence (*p*-value <0.10) endorsed by reporter assays, western blot, microarray profiling, and next-generation sequencing from miRTarBase v9.0[17]. We performed two complementary over-representation analyses with clusterProfiler [18]: (i) single-miRNA analyses of the effect of each miRNA on its target genes, and (ii) deregulated target genes corresponding to all differentially expressed miRNAs. Both analyses revealed which pathways were significantly enriched in the miRNA target genes. These analyses used the following collections in the Molecular Signatures Database: c5.go.bp from the Biological Process Gene Ontology [19], c2.reactome from the Reactome pathway database [20], and c2.kegg from the Kyoto Encyclopedia of Genes and Genomes pathway database [21]. Protein-protein interactions for the enriched genes were obtained using the *rba_string_interactions_network* function from the *rbioapi* package [22]. For interpretability in this manuscript, we highlighted lipid/atherosclerosis-related terms and filter keyword reporting to “cholesterol,” “HDL,” “atherosclerosis,” “vascular,” and “heart”. Analyses were conducted in R v. 4.2.2.

### 2.9. Data sharing statement

The data underlying this article are not publicly available due to ethical and legal restrictions related to participant privacy and informed consent. De-identified/pseudonymized data from the Girona Heart Registry (REGICOR) may be made available for collaborative research upon reasonable request and subject to review, ethics approval where applicable, and execution of a data transfer/use agreement. Requests should be directed to the REGICOR Study Group via the REGICOR <u>contact page</u>.

## 3. RESULTS

### 3.1. Study population

AMI cases were more likely to be men, older, had higher total and LDL cholesterol and triglycerides, lower HDL cholesterol, and presented a higher prevalence of hypercholesterolemia, hypertension, diabetes, and smoking habit (**Table 1**). Compared with the overall REGICOR population, the study subcohort had a slightly higher proportion of women and an older average age, and a lower prevalence of hypercholesterolemia and higher HDL cholesterol concentrations (**Supplementary Table 1**).

**Table 1.**
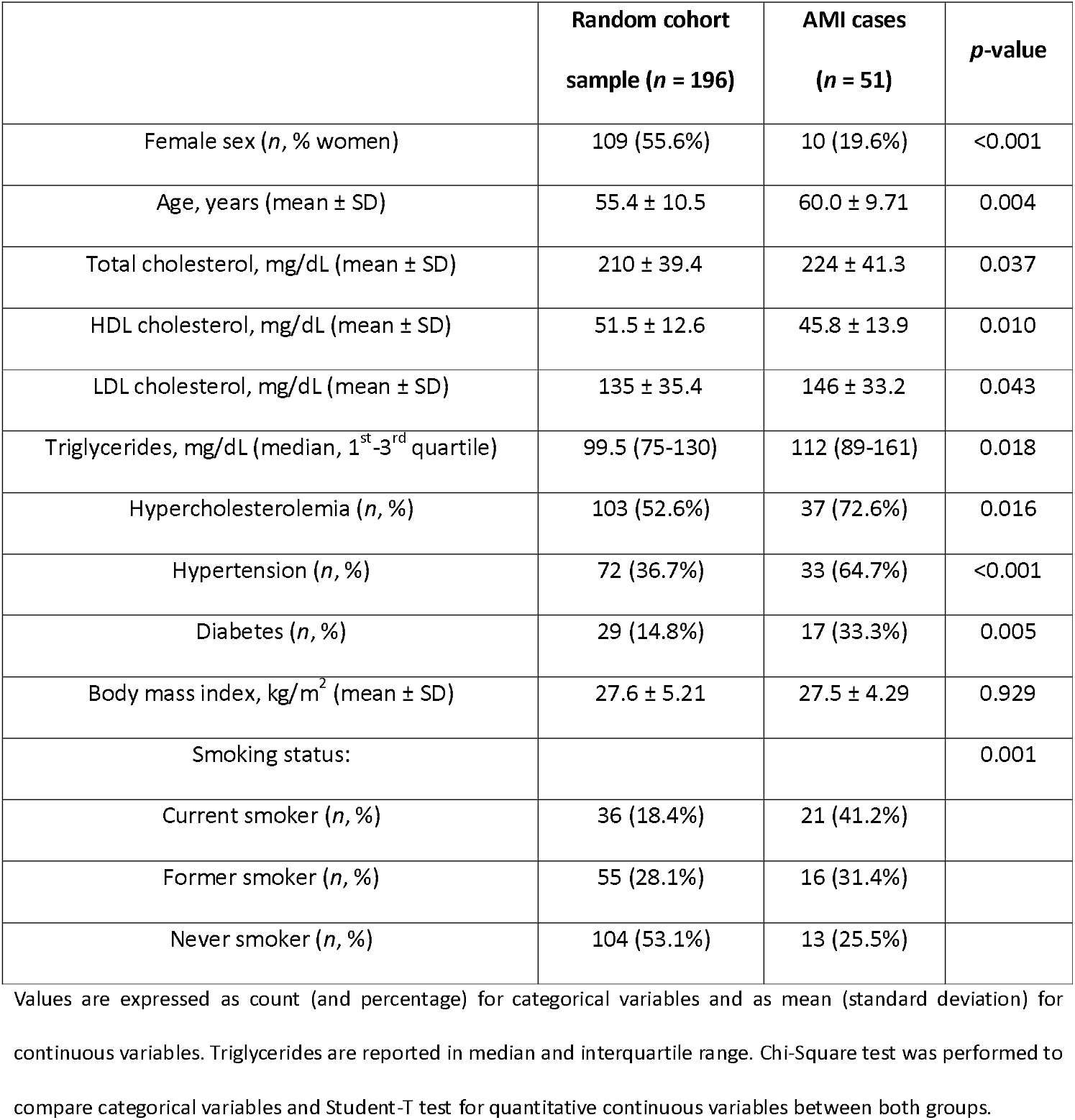
Study population characteristics.

### 3.2. HDL-miRNAs and incidence of acute myocardial infarction

As seen in **Figure 2**, levels of two HDL-miRNAs were associated with higher AMI incidence after multiple testing correction: miR-628-3p, HR 1.69, 95% Confidence Interval (CI) [1.30 to 2.19] and miR-28-3p, HR 1.58, 95% CI [1.21 to 2.06]. Nine other miRNAs were nominally linked to AMI incidence (*p*-value < 0.05), eight with higher AMI risk (miR-93-5p HR 1.87, 95% CI [1.16 to 3.02], miR-26b-5p HR 1.72, 95% CI [1.10 to 2.69], let-7f-5p HR 1.63, 95% CI [1.00 to 2.63], miR-106a-5p HR 1.49, 95% CI [1.04 to 2.12], let-7e-5p [HR [1.44, 95% CI [1.04 to 1.98], let-7a-5p HR 1.38, 95% CI [1.10 to 1.72], miR-126-3p [HR 1.37, 95% CI [1.02 to 1.85], and miR-15b-5p HR 1.21, 95% CI 1.00 to 1.47]) and one showing an inverse association (miR-361-5p [HR 0.69, 95% CI 0.48 to 1.00]). Nine other HDL-miRNAs showed tentative associations with AMI incidence (*p*-value < 0.1), six showing a direct association (miR-21-5p, miR-629-5p, let-7g-5p, let-7b-5p, miR-25-3p, and miR-195-5p) and three showing an inverse association (miR-1-3p, miR-4508, and miR-424-5p).

**Figure 2.**
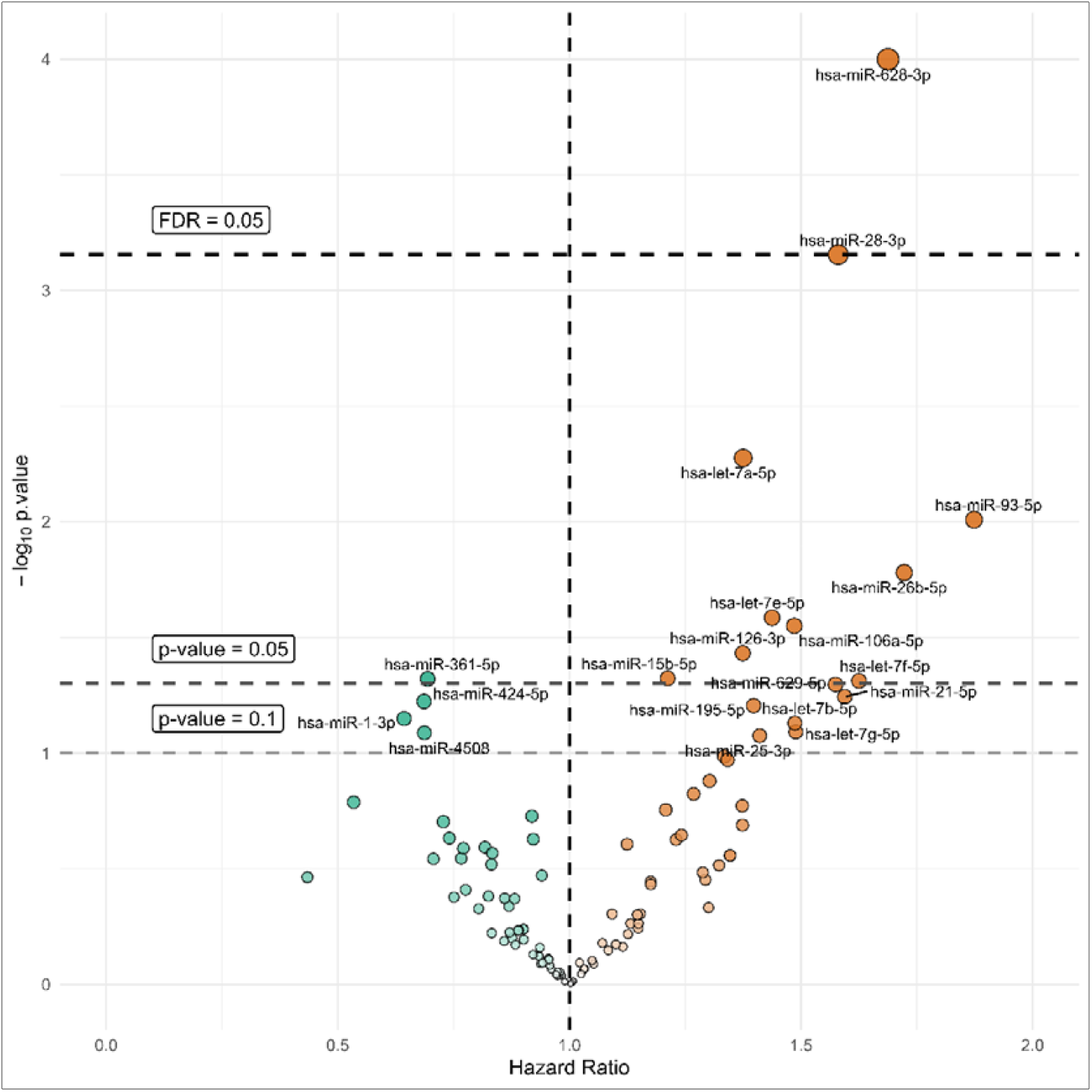
Volcano plot of the association between HDL-miRNA levels and incidence of AMI. The x-axis shows the hazard ratio from Cox models and the y-axis the corresponding −log_10_(p value). Horizontal dashed lines mark p = 0.10 and p = 0.05, and the thick dashed line the FDR threshold of 0.05.

### 3.3. Functional analysis in HDL-microRNAs: co-regulation and pathways

Principal component (PC) analyses of the selected HDL-miRNAs (**Supplementary Figures 1A-B**; loadings in **Supplementary Table 2**) revealed contribution to miRNAs expression variance. In AMI cases, dimension #1 (PC1) was explained by miR-93-5p and let-7 members (let-7f-5p, let-7b-5p, let-7g-5p) and capturing a 26.1% of variance, while dimension #2 (PC2) was dominated by miR-628-3p, miR-28-3p and miR-21-5p (signals that were negligible in controls). In controls, PC1 was again captured by miR-93-5p and let-7 family members (let-7f-5p, let-7b-5p, let-7g-5p), while PC2 variability was characterized by other miRNAs (let-7a-5p miR-26b-5p, let-7e-5p, miR-15b-5p).

Within-group correlation heatmaps (**Supplementary Figures 2A-B**) in AMI cases showed strong correlations (r>0.5) within the let-7 family and between let-7 members and miR-93-5p/miR-26b-5p, whilst in controls we found a sparse matrix with a few moderate links, largely within let-7 members.

HDL-miRNAs linked to AMI incidence targeted pathways involved in the regulation of cholesterol metabolism, particularly cholesterol efflux and transport (**Figure 3, Supplementary Table 3**). Single-miRNA over-representation analyses revealed that, in AMI cases, miR-26b-5p mapped to seven enriched processes driven by *ABCA1* and miR-126-3p connected to cholesterol efflux/transport through several validated targets (*SIRT1, NFKBIA, LRP6* or *ANXA2*) **(Supplementary Table 3)**.

**Figure 3.**
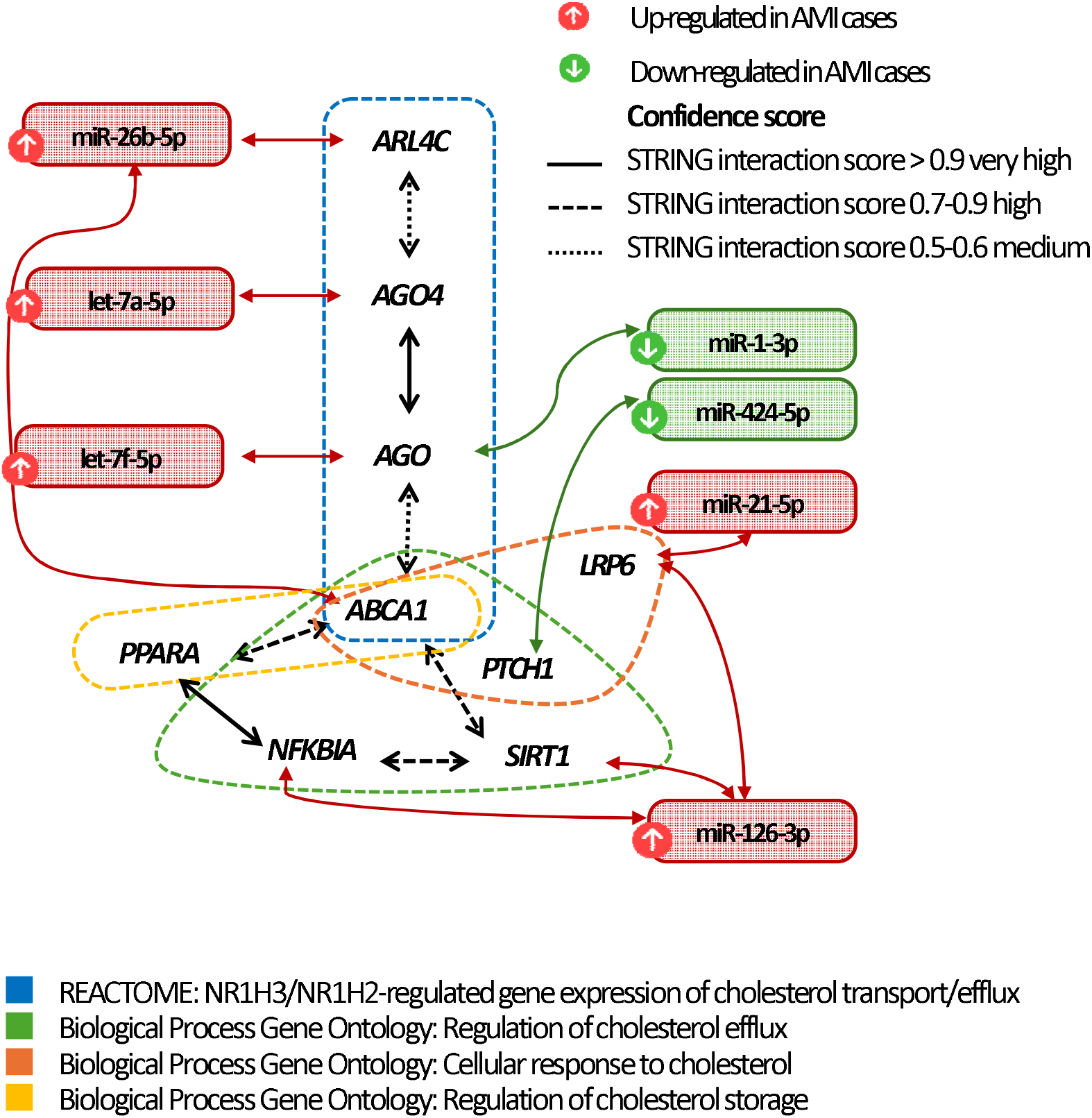
Network of AMI-linked HDL-miRNAs and deregulated targets within cholesterol-related pathways. The figure shows HDL-miRNAs significantly associated with incident AMI (FDR < 0.05) and their experimentally validated gene targets in enriched Reactome and Gene Ontology cholesterol-related processes. Red and green indicate miRNAs up- and down-regulated in AMI cases, and line styles reflect STRING interaction confidence.

## 4. DISCUSSION

No previous studies, to our knowledge, have reported a population-based untargeted study to show the role of HDL-miRNAs in AMI incidence. In our study two miRNAs (miR-628-3p and miR-28-3p) were associated with a higher incidence of AMI, nine more showed nominal associations (miR-93-5p, miR-26b-5p, miR-106a-5p, miR-126-3p, miR-15b-5p, miR-361-5p, let-7a-5p, let-7e-5p, and let-7f-5p) and functional analyses revealed roles in regulating cholesterol efflux and homeostasis in humans. We also captured higher variance and abundance of significant and strong correlations within the AMI group. Accordingly, in the principal component analysis, we observed a different contribution of miRNAs between controls and AMI cases group in PC2.

Two HDL-miRNAs were significantly associated with increased AMI incidence: miR-628-3p and miR-28-3p. To the best of our knowledge, this is the first report linking miR-628-3p, whether bound to HDL or not, to incident AMI, although circulating miR-628-3p levels are elevated in prevalent cases of several chronic and infectious diseases such as preeclampsia [23], certain adenocarcinomas [24] or viral infections [25,26]. miR-28-3p has already been linked to a greater mortality risk in acute coronary syndrome patients [27], and its circulating levels are higher in patients with pulmonary embolism [28], and some solid neoplasia [29,30]. To our knowledge, this is the first study that has linked miR-28-3p to higher AMI incidence, whether HDL-bound or freely circulating. In our data, miR-21-5p moderately correlated with miR-628-3p and miR-28-3p in AMI cases and was nominally associated with greater AMI incidence. In candidate miRNA studies, miR-21-5p levels have been linked to a greater incidence of myocardial infarction [9], complications of atherosclerotic disease[31], mortality after cardiogenic shock [32], and prevalence of type 2 diabetes with vascular complications [33] but, unexpectedly, to a lower risk of recurrent CVD events in individuals with prior AMI [34]. Altogether, these findings suggest the clinical utility of the HDL-bound miR-628-3p/miR-28-3p/miR-21-5p profile for AMI risk prediction in general population.

Another group of HDL-miRNAs centered on the let-7 family and related miRNAs was linked to increased AMI incidence in our data: let-7a-5p, let-7e-5p, let-7f-5p, miR-93-5p, and miR-26b-5p showed nominally significant direct associations with incident AMI (p < 0.05), while let-7b-5p and let-7g-5p were suggestively associated (0.05 < p < 0.1). All these molecules were highly intercorrelated, and several targeted key molecular genes in *ABCA1*-dependent cholesterol homeostasis pathways (miR-26b-5p, let-7a-5p, let-7f-5p). High levels of several of these miRNAs have been reported in patients with coronary disease (let-7b, let-7f, let-7g, miR-93-5p) [35–38] and in those with a history of cerebrovascular disease (let-7a-5p, let-7b-5p, let-7f-5p) [39]. Additionally, miR-26b-5p levels have been linked to differences in prognosis after myocardial infarction [40], or heart failure, and venous thromboembolism [40,41]. Mechanistically, it can inhibit ABCA1 expression by targeting LXR and ARL4C, thereby modulating cholesterol-related pathways relevant to atherosclerosis [42]. Together with previous evidence, our findings suggest a plausible co-regulatory network of HDL-miRNAs linked to higher AMI risk.

Some other independent HDL-miRNAs showed nominal associations with greater (miR-106a-5p, miR-126-3p, miR-15b-5p) or lower AMI incidence (miR-361-5p) in our data. Among them, miR-126-3p is the most clearly linked to CVD, since it showed a strong capacity to modulate cholesterol efflux/homeostasis targets in our data (*NFKBIA, SIRT1, LRP6 and ANXA2*) [43,44], and its circulating levels have been associated with a higher incidence of coronary disease [45], stroke and heart failure [46], and with higher levels of atherosclerotic coronary plaque in previous studies. Evidence for the others is more heterogenous; regarding miR-106a-5p, circulating levels have been inversely related to fatal AMI incidence in a nested case-control study with candidate genes in a Scandinavian general population [47]. To date, no study has examined associations between circulating miR-15b-5p and miR-361-5p and incident CVD, but both appear to be upregulated in coronary artery disease patients [48].

HDL particles are not passive cholesterol shuttles; they also can carry a regulatory small-RNA cargo that can be delivered to vascular and immune cells, altering gene expression in pathways central to atherogenesis[10]. We previously observed impaired HDL functionality in acute coronary syndrome patients in the same cohort [2], and others have shown that miRNAs modulate cholesterol efflux/homeostasis and vascular repair [49,50]. In the present proposal, we employed an untargeted analysis approach and HDL-enriched serum fractions allowing for unbiased identification of HDL-miRNA profiles. Together, these data support two complementary uses. First, HDL-miRNA signatures integrate information on HDL function and plaque biology that is not captured by other HDL-related characteristics, thereby offering additive value for risk stratification. This aligned with our observation that specific HDL-miRNAs track incident AMI beyond classic risk factors. Second, because miRNA networks are druggable, interventions that improve HDL function or composition could shift the HDL-miRNA cargo, and HDL-mimetic nanoparticles may be used to deliver miRNA inhibitors/mimics to atherosclerotic tissues. Collectively, HDL-miRNAs are both integrative biomarkers and actionable nodes in CVD prevention.

Our study presents several limitations. First, our analyses are observational and, despite extensive adjustment for available covariates, causal conclusions should not be drawn from our findings. Our analyses are also based on a single measurement of HDL-miRNAs at baseline, which does not allow assessment of temporal changes. Second, our results may have low reproducibility in other studies due to technical challenges and a reduced number of findings due to sample size constraints, the limited number of AMI events in a case-cohort design, and multiple testing correction. Although no external validation cohort is available due to the lack of currently suitable biological samples of comparable incident AMI cases, confirmation of our findings in independent populations is warranted and replication efforts in an independent sample set are being pursued as part of our ongoing research strategy. The use of apolipoprotein B-depleted serum as an HDL source is a faster approach that may better preserve sample integrity, but it also has its own limitations (potential contamination, as apolipoprotein B-depleted serum is primarily composed of HDL particles but also can contain lipoprotein-associated enzymes, extracellular vesicles, and albumin). Third, the limited amount of published evidence on HDL-miRNAs, which also heavily relies on *in silico* enrichment and candidate miRNA studies, hinders direct comparisons with similar works. Finally, generalizability may be limited to community-dwelling adults aged 35-75 from a Mediterranean country and may not extend to other age groups, ancestries or background.

### Translational perspective

High-density lipoproteins are no longer viewed only as cholesterol carriers, but as complex particles that integrate metabolic and inflammatory signals. The ability of HDL to transport microRNAs adds a new layer of functional versatility, potentially capturing the cardiometabolic state. In our population-based study, HDL-bound miR-628-3p and miR-28-3p were associated with incident acute myocardial infarction, and their validated targets (including ABCA1, SIRT1, NFKBIA and LRP6) mapped to pathways central to cholesterol efflux, lipid homeostasis and inflammation. These findings support HDL-miRNAs as circulating reporters that could complement traditional risk factors to refine AMI risk stratification and guide the intensity of preventive strategies. Building on prior work linking HDL functionality to myocardial infarction, our data point to HDL-miRNA–regulated pathways as plausible drivers of HDL-related biology. From a translational standpoint, synthetic HDL (HDL-mimetic) particles offer a controlled platform to probe and, potentially, modulate selected miRNA–target networks relevant to ASCVD.

## Data Availability

All data produced in the present study are available upon reasonable request to the authors

## Abbreviations

ABCA1: ATP-Binding Cassette Subfamily A Member 1
AMI: acute myocardial infarction
CVD: cardiovascular disease
HDL: high-density lipoprotein
HDL-miRNAs: HDL-bound microRNAs
HR: hazard ratio
miRNA: microRNA
REGICOR: Registre Gironí del Cor
CI: confidence interval
SD: standard deviation

## Acknowledgements

We thank Gemma Blanchart and Sònia Gaixas for their assistance in the project and the REGICOR participants and personnel for their collaboration. We thank CIBER de Fisiopatología de la Obesidad y Nutrición (CIBEROBN), CIBER de Diabetes y Enfermedades Metabólicas Asociadas (CIBERDEM), CIBER de Epidemiología y Salud Pública (CIBERESP), and CIBER de Enfermedades Cardiovasculares (CIBERCV); initiatives of Instituto de Salud Carlos III (Madrid, Spain), and financed by FEDER funds (CB06/03, CB07/08/0016, CB16/11/00229, CB16/11/00246).

## Author Contributions

Conceptualization: M.F. Methodology: J.H.-R., O.C., M.F. Formal analysis: J.H.-R. Investigation: J.H.-R., M.L., Á.H., P.B., D.M.-A., M.B., J.P., N.R., J.C.E.-G., D.C., O.K., A.C.-V., I.S., J.M., R.E., A.G., L.R., M.C.-S., D.M., M.H.H., A.S.-V., I.L. Resources: J.M., M.F. Data Curation: J.H.-R., M.L., P.B., D.M.-A., M.B., J.P., N.R., D.M. Writing - Original Draft: J.H.-R. Writing - Review & Editing: M.L., Á.H., P.B., D.M.-A., M.B., J.P., N.R., J.C.E.-G., D.C., O.K., A.C.-V., I.S., J.M., R.E., A.G., L.R., M.C.-S., D.M., M.H.H., I.R-D, O.C., M.F. Visualization: J.H.-R., Á.H. Project administration: J.M., M.F. Funding acquisition: M.F. M.F. had full access to all the data in the study and take responsibility for its integrity and the data analysis. All authors read and approved the final manuscript.

## Central Illustration

We used a population-based case-cohort design from REGICOR to test whether HDL-bound miRNAs predict incident AMI. HDL-miRNAs were quantified by whole-genome sequencing in apoB-depleted serum and analyzed with multivariable Cox models. Two HDL-miRNAs (miR-628-3p and miR-28-3p) were associated with AMI after multiple-testing correction, while additional HDL-miRNAs showed nominal associations and enriched targets in cholesterol efflux/homeostasis pathways. These results connect HDL regulatory cargo to future coronary events and motivate further clinical evaluation.

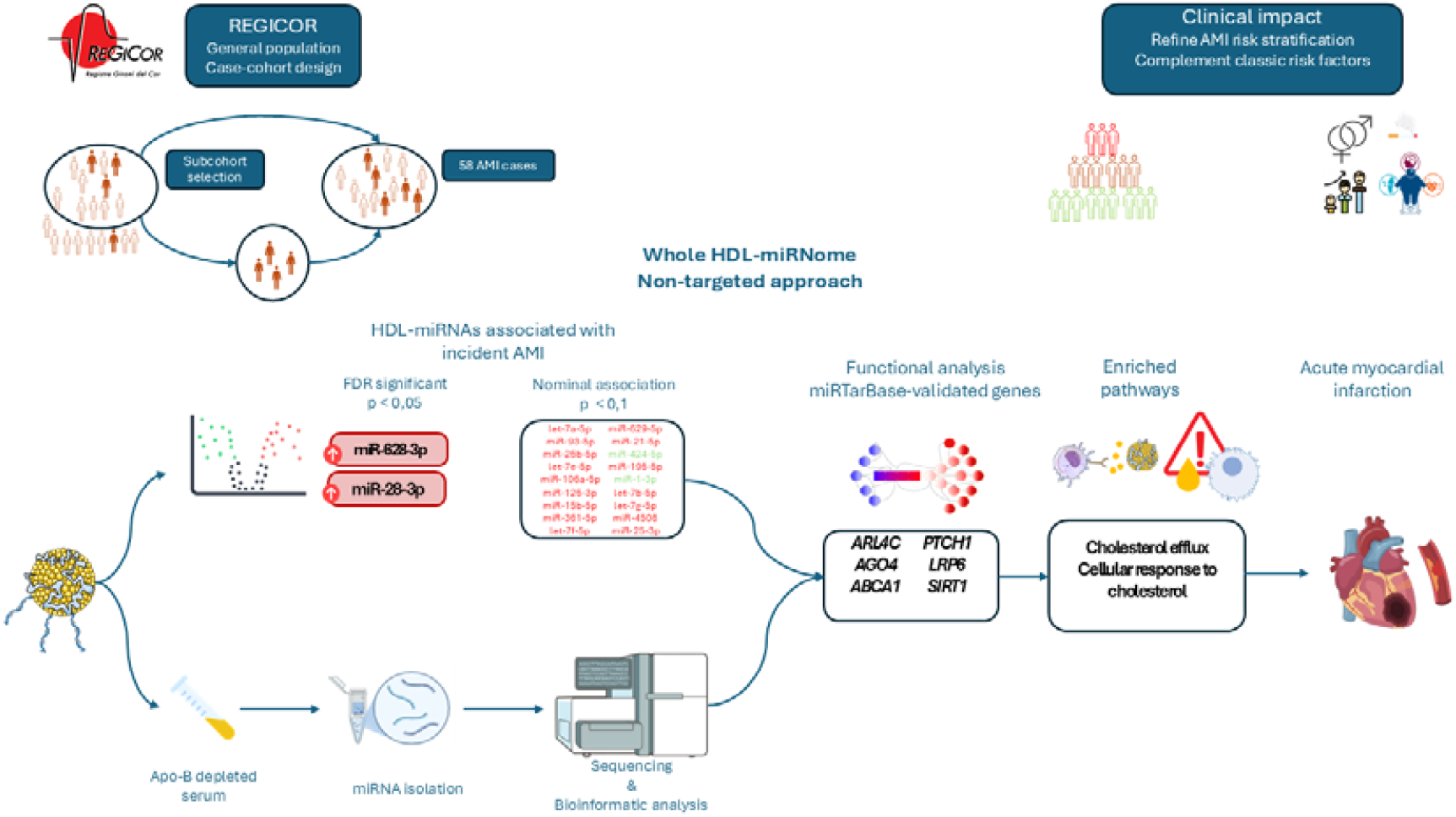

