## Supplementary material for "HDL-bound microRNAs modulating cholesterol efflux and homeostasis and incidence of acute myocardial infarction: A population-based case-cohort study"

**Supplementary Methods: Extraction, Sequencing, and Processing of miRNAs**

**A. Unique Molecular Identifiers Extraction and Preprocessing**

Unique Molecular Identifiers (UMIs) were extracted allowing for one mismatch with umitools version 1.1.2. After removing reads shorter than 15bp with Cutadapt v4.1, the total counts were reduced to ~3.7 million reads.

**B. Alignment to Reference**

Reads were aligned to miRBase v22.1 (with hairpin sequences converted to cDNA) using STAR v2.7.8a. Alignments were performed on a minimum of 15 matched bases (--outFilterMatchNmin 15) and a maximum 5% of mismatches (--outFilterMismatchNoverLmax 0.05) per alignment. No supplementary filters for matches and scores were applied (--outFilterScoreMinOverLread 0 --outFilterMatchNminOverLread 0).

**C. Quantification**

The quantification was generated using the Binary Alignment Map (BAM, -R BAM) produced by featureCounts function in the Subread package v2.0.3 followed by umi_tools count with unique method. Multimapping reads were accounted for (-M --fraction), and a minimum of 85% overlap with the miRNA feature was required (--fracOverlapFeature 0.85).

**D. Filtering and Normalization**

Lowly expressed miRNAs, defined as less than 3 counts in at least 50 samples, were excluded from further analysis, resulting in 111 miRs. Raw library size differences between samples were approached following the weighted “trimmed mean method” TMM ^1^ implemented in the edgeR package ^2^.

**E. Quality Control**

There was no clear pattern of clustering or batch effect either, except for one (number 5), which has been employed in the adjustment procedure. The detected volume effect corresponds also to the samples with the lowest counts that were removed. Additionally, samples with less than 1.000 total counts were also excluded completing 246 total samples.

**Supplementary tables**

**Supplementary table 1 -** Comparison of Baseline Characteristics Between Study Subcohort and Overall REGICOR Population.

|  | **Random subcohort (n = 196)** | **REGICOR population (n = 5404)** | **p-value** |
| --- | --- | --- | --- |
| Sex (n, % women) | 109 (55.6%) | 2915 (54%) | 0.001 |
| Age, years (mean ± SD) | 55.4 (10.5) | 53.7 (10.9) | 0.028 |
| Weight, Kg (mean ± SD) | 73.7 (15.9) | 73.1 (14.3) | 0.592 |
| BMI, kg/m^2^ (mean ± SD) | 27.6 (5.21) | 27.2 (4.63) | 0.309 |
| Hypertension (n, %) | 72 (36.7%) | 2226 (41.3%) | 0.231 |
| Diabetes (n, %) | 29 (14.8%) | 700 (13%) | 0.522 |
| Hypercholesterolemia (n, %) | 103 (52.6%) | 3525 (65.3%) | <0.001 |
| Total cholesterol, mg/dL (mean ± SD) | 210.4 (39.4) | 211.4 (41.8) | 0.709 |
| LDL cholesterol, mg/dL (mean ± SD) | 134.9 (35.4) | 136.5 (36.8) | 0.554 |
| HDL cholesterol, mg/dL (mean ± SD) | 51.6 (12.6) | 54.2 (15.2) | 0.008 |
| Triglycerides, mg/dL (median, 1^st^-3^rd^ quartile) | 99.5 [75 - 130] | 93 [68 - 132] | 0.146 |
| Smoking status (n, %) |  |  |  |
| Regularly | 36 (18.4%) | 1174 (21.7%) | 0.300 |
| Former smoker | 55 (28.1%) | 1435 (26.6%) | 0.700 |
| Never smoker | 104 (53.1%) | 2740 (50.7%) | 0.566 |

BMI: body mass index (kg/m^2^); Values are expressed as count (percentage) for categorical variables and as mean (standard deviation) for continuous variables. Triglycerides are reported in median and interquartile range. Chi-Square test was performed to compare categorical variables and Student-T test for quantitative continuous variables between both groups.

**Supplementary table 2 –** Principal Component Loadings of miRNAs in Controls and AMI Cases. Bold indicates principal component loadings greater than 0.35.

| **Controls** | **PC1** | **PC2** | **AMI Cases** | **PC1** | **PC2** |
| --- | --- | --- | --- | --- | --- |
| hsa-miR-628-3p | 0.048 | 0.208 | hsa-miR-628-3p | 0.017 | **0.486** |
| hsa-miR-28-3p | -0.108 | 0.044 | hsa-miR-28-3p | 0.077 | **0.456** |
| hsa-let-7a-5p | 0.218 | **-0.388** | hsa-let-7a-5p | 0.297 | -0.11 |
| hsa-miR-93-5p | **0.401** | 0.209 | hsa-miR-93-5p | **0.397** | -0.021 |
| hsa-miR-26b-5p | 0.213 | **0.376** | hsa-miR-26b-5p | 0.293 | 0.039 |
| hsa-let-7e-5p | 0.201 | **-0.391** | hsa-let-7e-5p | 0.094 | -0.181 |
| hsa-miR-106a-5p | 0.202 | 0.066 | hsa-miR-106a-5p | 0.221 | -0.07 |
| hsa-miR-126-3p | -0.07 | 0.258 | hsa-miR-126-3p | -0.082 | -0.047 |
| hsa-miR-15b-5p | 0.073 | **0.356** | hsa-miR-15b-5p | -0.002 | 0.098 |
| hsa-miR-361-5p | -0.055 | 0.237 | hsa-miR-361-5p | -0.126 | 0.029 |
| hsa-let-7f-5p | **0.431** | -0.018 | hsa-let-7f-5p | 0.347 | -0.109 |
| hsa-miR-629-5p | -0.027 | 0.07 | hsa-miR-629-5p | -0.185 | -0.105 |
| hsa-miR-21-5p | -0.094 | 0.281 | hsa-miR-21-5p | 0.067 | **0.523** |
| hsa-miR-424-5p | -0.101 | 0.222 | hsa-miR-424-5p | 0.052 | -0.011 |
| hsa-miR-195-5p | 0.203 | 0.13 | hsa-miR-195-5p | 0.249 | 0.112 |
| hsa-miR-1-3p | -0.082 | 0.067 | hsa-miR-1-3p | -0.188 | -0.301 |
| hsa-let-7b-5p | **0.415** | -0.091 | hsa-let-7b-5p | **0.384** | -0.101 |
| hsa-let-7g-5p | **0.452** | 0.095 | hsa-let-7g-5p | **0.401** | -0.046 |
| hsa-miR-4508 | -0.048 | -0.108 | hsa-miR-4508 | 0.107 | -0.27 |
| hsa-miR-25-3p | 0.074 | 0.172 | hsa-miR-25-3p | 0.05 | 0.085 |

^a^PC: principal components.

**Supplementary Table 2** - Single miRNA Enrichment Analysis of Cholesterol-Related Pathways in Control and Case Groups.

| **Control sample group** | | | | | |
| --- | --- | --- | --- | --- | --- |
| **Biological process** | **p-value** | **FDR** | **q-value** | **geneID** | **miRNA** |
| REACTOME_REGULATION_OF_CHOLESTEROL_BIOSYNTHESIS_BY_SREBP_SREBF | 0,024 | 0,050 | 0,020 | SP1/FASN | hsa-miR-1-3p |
| **Myocardial infarction cases group** | | | | | |
| **Biological process** | **p-value** | **FDR** | **q-value** | **geneID** | **miRNA** |
| GOBP_VESICLE_MEDIATED_CHOLESTEROL_TRANSPORT | 0,011 | 0,046 | **0,019** | *SYT7* | hsa-let-7e-5p |
| REACTOME_CHOLESTEROL_BIOSYNTHESIS | 0,028 | 0,082 | **0,043** | *FDPS* | hsa-let-7f-5p |
| REACTOME_NR1H3_NR1H2_REGULATE_GENE_EXPRESSION_LINKED_TO_CHOLESTEROL_  TRANSPORT_AND_EFFLUX | 0,053 | 0,082 | **0,043** | *AGO1* | hsa-let-7f-5p |
| REACTOME_REGULATION_OF_CHOLESTEROL_BIOSYNTHESIS_BY_SREBP_SREBF | 0,073 | 0,092 | **0,048** | *FDPS* | hsa-let-7f-5p |
| REACTOME_HDL_ASSEMBLY | 0,018 | 0,068 | **0,037** | *ABCA1* | hsa-miR-26b-5p |
| REACTOME_NR1H3_NR1H2_REGULATE_GENE_EXPRESSION_LINKED_TO_CHOLESTEROL_  TRANSPORT_AND_EFFLUX | 0,004 | 0,022 | **0,012** | *ABCA1/ARL4C* | hsa-miR-26b-5p |
| GOBP_NEGATIVE_REGULATION_OF_CHOLESTEROL_STORAGE | 0,024 | 0,077 | **0,042** | *ABCA1* | hsa-miR-26b-5p |
| GOBP_REVERSE_CHOLESTEROL_TRANSPORT | 0,030 | 0,078 | **0,042** | *ABCA1* | hsa-miR-26b-5p |
| GOBP_REGULATION_OF_CHOLESTEROL_STORAGE | 0,039 | 0,086 | **0,047** | *ABCA1* | hsa-miR-26b-5p |
| GOBP_CHOLESTEROL_STORAGE | 0,054 | 0,093 | **0,050** | *ABCA1* | hsa-miR-26b-5p |
| GOBP_POSITIVE_REGULATION_OF_CHOLESTEROL_EFFLUX | 0,057 | 0,093 | **0,050** | *ABCA1* | hsa-miR-26b-5p |
| REACTOME_NR1H2_NR1H3_REGULATE_GENE_EXPRESSION_TO_LIMIT_CHOLESTEROL_  UPTAKE | 0,015 | 0,045 | **0,016** | *MYLIP* | hsa-mirR-106a-5p |
| GOBP_POSITIVE_REGULATION_OF_CHOLESTEROL_EFFLUX | 0,002 | 0,008 | **0,003** | *SIRT1/NFKBIA* | hsa-miR-126-3p |
| GOBP_REGULATION_OF_CHOLESTEROL_EFFLUX | 0,004 | 0,014 | **0,005** | *SIRT1/NFKBIA* | hsa-miR-126-3p |
| GOBP_CHOLESTEROL_EFFLUX | 0,010 | 0,033 | **0,012** | *SIRT1/NFKBIA* | hsa-miR-126-3p |
| GOBP_RECEPTOR_MEDIATED_ENDOCYTOSIS_INVOLVED_IN_CHOLESTEROL_TRANSPORT | 0,018 | 0,045 | **0,016** | *ANXA2* | hsa-miR-126-3p |
| GOBP_CELLULAR_RESPONSE_TO_CHOLESTEROL | 0,048 | 0,074 | **0,027** | *LRP6* | hsa-miR-126-3p |
| REACTOME_REGULATION_OF_CHOLESTEROL_BIOSYNTHESIS_BY_SREBP_SREBF | 0,102 | 0,123 | **0,045** | *MTF1* | hsa-miR-25-3p |

**Supplementary Figures**

**Supplementary Figure 1A and 1B -** Principal Component Analysis Biplot in Controls and Cases

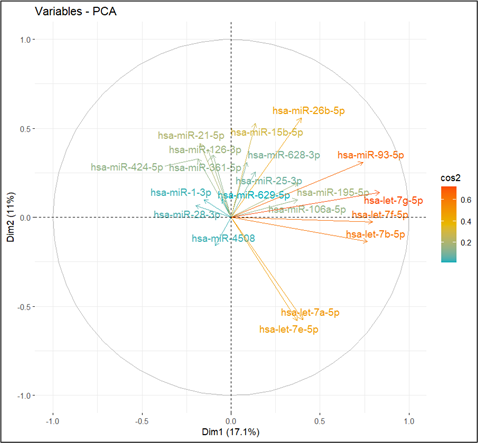

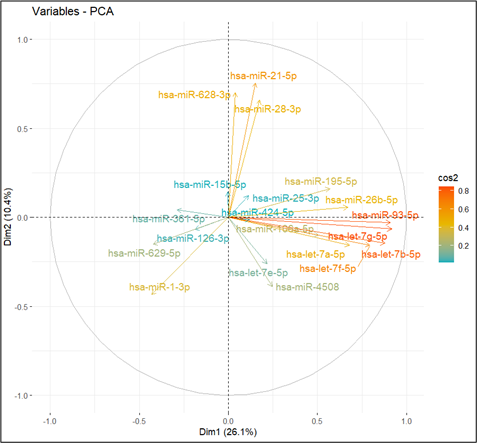

**Supplementary figure 2A and 2B -** Pairwise Correlation Analysis of Selected miRNAs in Controls and Cases. The heatmaps display Pearson’s correlation coefficients between selected HDL-bound miRNAs significantly associated after multiple-testing correction (FDR-adjusted p < 0.05).

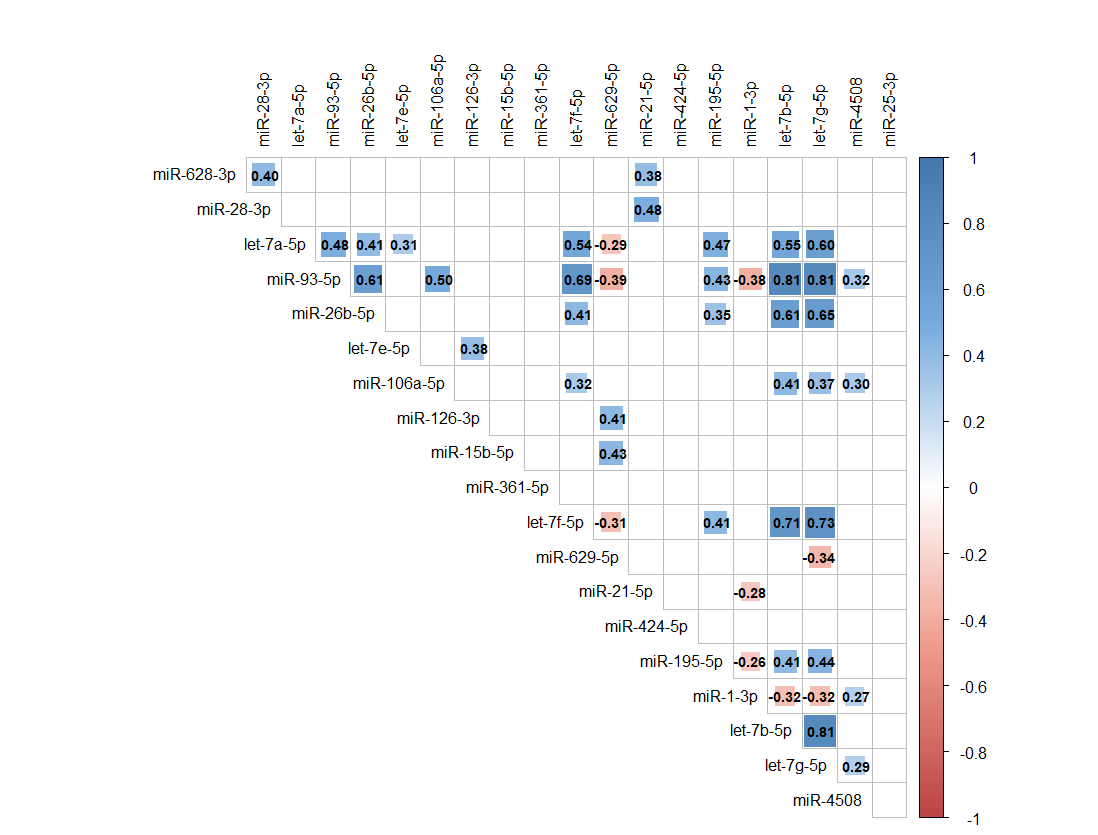

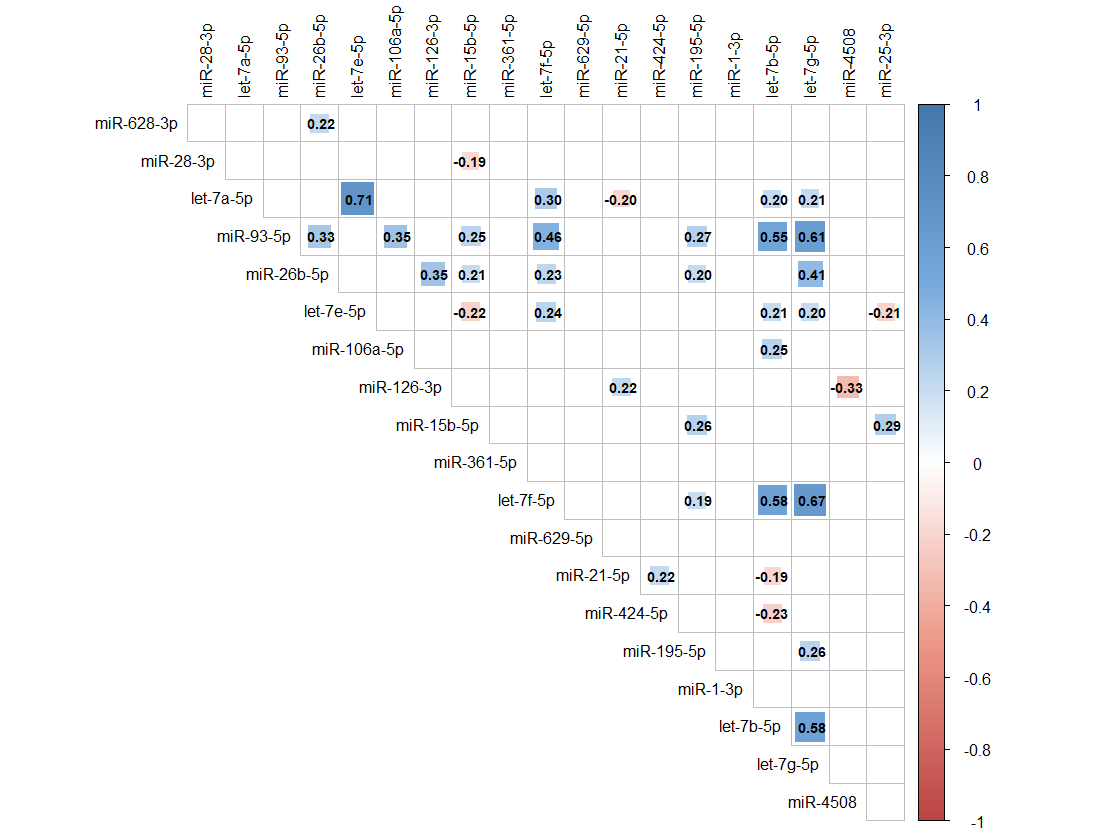
