## Supplementary tables for "HDL-bound microRNAs modulating cholesterol efflux and homeostasis and incidence of acute myocardial infarction: A population-based case-cohort study"

**Table 1.** Study population characteristics.

|  | **Random cohort**  **sample (*n* = 196)** | **AMI cases (*n* = 51)** | ***p*-value** |
| --- | --- | --- | --- |
| Female sex (*n*, % women) | 109 (55.6%) | 10 (19.6%) | <0.001 |
| Age, years (mean ± SD) | 55.4 ± 10.5 | 60.0 ± 9.71 | 0.004 |
| Total cholesterol, mg/dL (mean ± SD) | 210 ± 39.4 | 224 ± 41.3 | 0.037 |
| HDL cholesterol, mg/dL (mean ± SD) | 51.5 ± 12.6 | 45.8 ± 13.9 | 0.010 |
| LDL cholesterol, mg/dL (mean ± SD) | 135 ± 35.4 | 146 ± 33.2 | 0.043 |
| Triglycerides, mg/dL (median, 1^st^-3^rd^ quartile) | 99.5 (75-130) | 112 (89-161) | 0.018 |
| Hypercholesterolemia (*n*, %) | 103 (52.6%) | 37 (72.6%) | 0.016 |
| Hypertension (*n*, %) | 72 (36.7%) | 33 (64.7%) | <0.001 |
| Diabetes (*n*, %) | 29 (14.8%) | 17 (33.3%) | 0.005 |
| BMI, kg/m^2^ (mean ± SD) | 27.6 ± 5.21 | 27.5 ± 4.29 | 0.929 |
| Smoking status: |  |  | 0.001 |
| Current smoker (*n*, %) | 36 (18.4%) | 21 (41.2%) |  |
| Former smoker (*n*, %) | 55 (28.1%) | 16 (31.4%) |  |
| Never smoker (*n*, %) | 104 (53.1%) | 13 (25.5%) |  |

Values are expressed as count (and percentage) for categorical variables and as mean (standard deviation) for continuous variables. Triglycerides are reported in median and interquartile range. Chi-Square test was performed to compare categorical variables and Student-T test for quantitative continuous variables between both groups.

**Supplementary table 1 -** Strengthening the Reporting of Observational Studies (STROBE) Statement checklist of items that should be included in reports of observational studies

|  | | Item No | | Recommendation | Page  No |
| --- | --- | --- | --- | --- | --- |
| **Title and abstract** | | 1 | | (*a*) Indicate the study’s design with a commonly used term in the title or the abstract | 1 |
|  |  |  |  | (*b*) Provide in the abstract an informative and balanced summary of what was done and what was found | 4 |
| Introduction | | | | | |
| Background/rationale | | 2 | | Explain the scientific background and rationale for the investigation being reported | 7 |
| Objectives | | 3 | | State specific objectives, including any prespecified hypotheses | 7 |
| Methods | | | | | |
| Study design | | 4 | | Present key elements of study design early in the paper | 8 |
| Setting | | 5 | | Describe the setting, locations, and relevant dates, including periods of recruitment, exposure, follow-up, and data collection | 8 |
| Participants | | 6 | | (*a*) *Cohort study*—Give the eligibility criteria, and the sources and methods of selection of participants. Describe methods of follow-up  *Case-control study*—Give the eligibility criteria, and the sources and methods of case ascertainment and control selection. Give the rationale for the choice of cases and controls  *Cross-sectional study*—Give the eligibility criteria, and the sources and methods of selection of participants | 8 |
|  |  |  |  | (*b*) *Cohort study*—For matched studies, give matching criteria and number of exposed and unexposed  *Case-control study*—For matched studies, give matching criteria and the number of controls per case | - |
| Variables | | 7 | | Clearly define all outcomes, exposures, predictors, potential confounders, and effect modifiers. Give diagnostic criteria, if applicable | 9, 10 |
| Data sources/ measurement | | 8* | | For each variable of interest, give sources of data and details of methods of assessment (measurement). Describe comparability of assessment methods if there is more than one group | 9, Supplementary  methods |
| Bias | | 9 | | Describe any efforts to address potential sources of bias | 10 |
| Study size | | 10 | | Explain how the study size was arrived at | 10, 11 |
| Quantitative variables | | 11 | | Explain how quantitative variables were handled in the analyses. If applicable, describe which groupings were chosen and why | 11 |
| Statistical methods | | 12 | | (*a*) Describe all statistical methods, including those used to control for confounding | 11 |
|  |  |  |  | (*b*) Describe any methods used to examine subgroups and interactions | - |
|  |  |  |  | (*c*) Explain how missing data were addressed | 11 |
|  |  |  |  | (*d*) *Cohort study*—If applicable, explain how loss to follow-up was addressed  *Case-control study*—If applicable, explain how matching of cases and controls was addressed  *Cross-sectional study*—If applicable, describe analytical methods taking account of sampling strategy | - |
|  |  |  |  | (*e*) Describe any sensitivity analyses |  |
| Results | | | | | |
| Participants | 13* | | (a) Report numbers of individuals at each stage of study—eg numbers potentially eligible, examined for eligibility, confirmed eligible, included in the study, completing follow-up, and analysed | | 13 |
|  |  |  | (b) Give reasons for non-participation at each stage | | - |
|  |  |  | (c) Consider use of a flow diagram | | 8 |
| Descriptive data | 14* | | (a) Give characteristics of study participants (eg demographic, clinical, social) and information on exposures and potential confounders | | 13 |
|  |  |  | (b) Indicate number of participants with missing data for each variable of interest | | - |
|  |  |  | (c) *Cohort study*—Summarise follow-up time (eg, average and total amount) | | 8 |
| Outcome data | 15* | | *Cohort study*—Report numbers of outcome events or summary measures over time | | 8 |
|  |  |  | *Case-control study—*Report numbers in each exposure category, or summary measures of exposure | | - |
|  |  |  | *Cross-sectional study—*Report numbers of outcome events or summary measures | | - |
| Main results | 16 | | (*a*) Give unadjusted estimates and, if applicable, confounder-adjusted estimates and their precision (eg, 95% confidence interval). Make clear which confounders were adjusted for and why they were included | | 14 |
|  |  |  | (*b*) Report category boundaries when continuous variables were categorized | | 14 |
|  |  |  | (*c*) If relevant, consider translating estimates of relative risk into absolute risk for a meaningful time period | | - |
| Other analyses | 17 | | Report other analyses done—eg analyses of subgroups and interactions, and sensitivity analyses | | 15 |
| Discussion | | | | | |
| Key results | 18 | | Summarize key results with reference to study objectives | | 17 |
| Limitations | 19 | | Discuss limitations of the study, taking into account sources of potential bias or imprecision. Discuss both direction and magnitude of any potential bias | | 19 |
| Interpretation | 20 | | Give a cautious overall interpretation of results considering objectives, limitations, multiplicity of analyses, results from similar studies, and other relevant evidence | | 17, 18, 19 |
| Generalizability | 21 | | Discuss the generalizability (external validity) of the study results | | 21 |
| Other information | | | | | |
| Funding | 22 | | Give the source of funding and the role of the funders for the present study and, if applicable, for the original study on which the present article is based | | 22 |

^a^PC: principal component
